# Social capital, urbanization level, and COVID-19 vaccination uptake in the United States: A national level analysis

**DOI:** 10.1101/2022.03.04.22271917

**Authors:** Shan Qiao, Zhenlong Li, Jiajia Zhang, Xiaowen Sun, Camryn Garrett, Xiaoming Li

**Affiliations:** Department of Health Promotion, Education, and Behavior, Arnold School of Public Health, University of South Carolina; Department of Geography, University of South Carolina; Department of Epidemiology and Biostatistics, Arnold School of Public Health, University of South Carolina

**Keywords:** COVID-19, Vaccination, Social Capital, Urbanization level, United States

## Abstract

Vaccination remains the most promising mitigation strategy for the COVID-19 pandemic. However, existing literature shows significant disparities in vaccination uptake in the United States. Using publicly available national-level data, we aimed to explore if county-level social capital can further explain disparities in vaccination uptake rate adjusting for demographic and social determinants of health (SDOH) variables; and if association between social capital and vaccination uptake may vary by urbanization level. Bivariate analyses and hierarchical multivariable quasi-binomial regression analysis were conducted, then the regression analysis was stratified by urban-rural status. The current study suggests that social capital contributes significantly to the disparities of vaccination uptake in the US. The results of stratification analysis show common predictors of vaccine uptake but also suggest various patterns based on urbanization level regarding the associations of COVID-19 vaccination uptake with SDOH and social capital factors. The study provides a new perspective to address disparities in vaccination uptake through fostering social capital within communities, which may inform tailored public health intervention efforts in enhancing social capital and promoting vaccination uptake.

## INTRODUCTION

High uptake of the COVID-19 vaccine is among the most promising strategies to reduce the burden of disease and control the pandemic [1]. Efficacious COVID-19 vaccines have been available and administered in the United States (US) since December 2020. [2]However, only 60.8% of US population have been fully vaccinated as of December 13, 2021[3] with notable disparities in vaccine uptake [4-6]. Per CDC vaccine administration data (12/14/2020-5/1/2021), vaccination coverage was lower in counties with high social vulnerability than in counties with low social vulnerability. This disparity was especially evident in large, fringe metropolitan (suburban) and nonmetropolitan counties [7]. As such, social factors are found to exacerbate the vulnerability of communities when stratified by demographic features (e.g., racial minority populations and age)[7]. Overall, vaccine uptake rates are lower in counties with higher percentages of older adults with a race/ethnicity other than non-Hispanic White, those without a computer or internet access, those living in poverty, and those living alone [8].

In addition to examining demographic characteristics [4,5] and structural-level factors (e.g., the social determinants of health [SDOH] and the social vulnerability index [SVI]) [6-11], research has increasingly turned its attention to the role of community-level factors (e.g., social capital) in disparities of vaccine uptake. Social capital is defined as the networks of relationships among people who live and work in a particular society which enables that society to function effectively [9]. Within a community where there exists trust and cooperation, a concentration of resources enables individuals to collaborate when faced with situations that call for collective action [10]. Despite the debates surrounding the concept of social capital over the past two decades, social capital represents a unique synthesis of research on social cohesion, income inequality, and social support. It is conceptualized as assets of places and communities, a contextual-level property, that is composed of cognitive domains (perceptions of trust, community beliefs and norms) and network domains (ties of friends, social participation, network-based resources, and social support) [11]. Early literature on social capital and health examines whether social capital mediates the association between income inequality and health. Recent studies focus primarily on inequalities in social capital and its contribution to health inequalities and disparities rather than their interplay [12].

The role of social capital on health may vary by urbanization level. Rural and urban settings differ in terms of population size, segregation of communities by social status (i.e., financial, educational, occupational, etc.), cohesion of community values, and the strength of geographic and non-geographic networks [7 13-15]. Geographic networks are groups of individuals who are united due to geographical proximity (i.e., neighborhoods, schools, workplaces, etc.), whereas non-geographic networks are groups of individuals who unite under a shared purpose that does not arise from geographical proximity (i.e., religious organizations, activist groups, support groups, etc.)[15]. The characteristics of geographic and non-geographic networks are shaped by spatial distribution, geographic distance, and transportation availability where communities exist within various statuses along the urbanization continuum [16]. These networks and their characteristics are likely to influence the ways in which community members contextualize social beliefs and norms as well as obtain resources and social support [16]. For example, certain domains of social capital, such as social cohesion, may affect individuals’ health-related behaviors differently in a metropolitan area as compared to a rural area because urban communities are more heterogeneous, and individuals are less likely to be influenced by social norms. Additionally, individuals living in a metropolitan area may have access to multiple non-social networks that provide social support and resources that extend beyond geographic networks or communities [17].

Within the context of vaccine uptake, existing literature suggests that social capital was associated with state-level vaccination uptake in the US during the 2009 H1N1 pandemic [18]. Social capital is understood to facilitate vaccine uptake within communities through enhancing trust, promoting altruism, and providing social support. Firstly, residing in a community with high levels of generalized trust is likely to be associated with immunization where trust in government and health institutions influences risk perceptions and health behaviors [19]. Those with high trust in government are more likely to support governmental policies regarding vaccination [19]. People who trust in health institutions are more likely to seek health care and ensure that they receive adequate treatment [20]. Institutional trust has been found to be significantly associated with acceptance of the Covid-19 vaccine [19]. Secondly, social capital is believed to foster altruism, which affects individual vaccination decision making. Vaccinating a society against communicable diseases can be characterized as a collective action, because doing so fosters herd immunity against the disease [15]. Eligible individuals may choose to contribute to herd immunity to protect the general other as well as those within their communities [15 18]. Social capital facilitates cooperation for altruistic collective action due to the influence of strong norms, enforcing acceptable behavior that tends to increase levels of cooperation, which may affect immunization decisions and reduce the prevalence of the reliance of individuals on herd immunity free-riding strategies (i.e., the individual benefits from herd immunity without contributing to it, in this case through vaccination) [18]. Finally, communities with high levels of social capital have access to more resources to overcome logistic challenges and barriers that hinder vaccination uptake such as transportation and vaccination appointment availability. For example, local health organizations, non-governmental organizations, and neighborhood committees may facilitate addressing these issues by offering patient navigation services, mobile vaccination clinics, and other health fair events to promote vaccine uptake, but may differ between communities due to levels of social capital [21].

While there is evidence of the association between social capital and vaccination uptake, few studies have examined how social capital level is related to rates of COVID-19 vaccination uptake at the county-level in the United States when controlling for demographic and SDOH variables. Additionally, although there are rationales to support that the influence of social capital on vaccine uptake may vary by urbanization level, there are limited data testing this hypothesis. Therefore, based on national data, the current study aims to answer two main questions: (1) Is social capital associated with vaccination uptake, when controlling for demographic characteristics and SDOH variables? and (2) Does the effect of social capital on vaccination uptake vary by urbanization level?

## METHODS

### Data sources

Data utilized in the current study come from five publicly available dataset sources (Table 1). Vaccination rates were extracted from CDC’s United States county and local estimates for vaccine hesitancy as of 12/23/2021 [22]. Demographic information was compiled from the U.S. Census American Community Survey (ACS), which includes 5-year population estimates (2015-2019) [23]. Variables representing SDOH were extracted from Emory University’s AIDSVu 2018 County SODH data [24]. Social capital variables were compiled utilizing data from the Social Capital Index Project aimed to understand the geography of social capital in America [25]. Finally, urbanization level was assessed using data from the CDC’s National Center for Health Statistics (NCHS) 2013 Rural Classification Scheme for Counties in the United States [26].

**Table 1.** Selected variables and data source.

| Category | Variables | Source |
| --- | --- | --- |
| Vaccination rate | % Adults fully vaccinated against COVID-19 (as of 6/10/21) | CDC Vaccine Hesitancy for COVID-19: County and local estimates<br>https://data.cdc.gov/Vaccinations/Vaccine-Hesitancy-for-COVID-19-County-and-local-es/q9mh-h2tw |
| Demographics | % Female<br>% Hispanic<br>% non-Hispanic Asian<br>% non-Hispanic Black<br>% non-Hispanic Native Hawaiian/Pacific Islander | US Census American Community Survey (ACS) 5-year estimate (2014-2018)<br>https://www.census.gov/newsroom/press-releases/2019/acs-5-year.html |
| Social Determinant of Health (SDOH) | % Living in Poverty<br>% High School Education<br>% Uninsured<br>% Unemployed<br>% Living with Severe Housing Cost Burden<br>Median Household Income<br>Gini Coefficient | AIDSVu 2018 County Social Determinants of Health.<br>https://aidsvu.org/ |
| Social Capital | % Births to unmarried women<br>% Women currently married<br>% Children with single parent<br>Non-religious non-profit organizations per 1000<br>Religious congregations per 1000<br>Presidential election voting rate 2012 & 2016<br>Mail-back census response rate<br>Violent Crimes per 100000 | Social Capital Index Project: The Geography of Social Capital in America.<br>https://www.lee.senate.gov/scp-index |
| Urban/Rural | 2013 urbanization code | CDC National Center for Health Statistics (NCHS) Rural Classification Scheme for Counties (2013)<br>https://www.cdc.gov/nchs/data_access/urban_rural.htm |

### Measures

#### Vaccination uptake rate

The county-level vaccine uptake rate was percent of adults (aged ≥18 years) who are fully vaccinated with any Food and Drug Administration (FDA)-authorized COVID-19 vaccine (have had the second dose of a two-dose vaccine or one dose of a single-dose vaccine) based on the jurisdiction and county where the recipient resides [27].

#### Demographic characteristics

Population demographic characteristics including gender, race, and ethnicity were aggregated to county-level. Specifically, the variables included percentage of female, percentage of Hispanic, percentage of non-Hispanic Asian, percentage of non-Hispanic Black, and percentage of non-Hispanic Native Hawaiian/Pacific Islander.

#### Social determinants of health (SDOH)

The SDOH variables included measures of poverty (percent of population living below federal poverty line)[28], high school education (percent of population with a high school degree or equivalent)[29], median household income[28], Gini Coefficient (a measure of income inequality with 0 reflecting complete equality and 1 reflecting complete inequality)[30 31], and percentage of population without health insurance [32].

#### Social capital

The Social Capital Index from the Social Capital Index Project in the United States was employed to estimate social capital. This index is composed of indicators in multilevel domains including family unity (e.g., births in past year to women who were unmarried and children living with a single parent), community health (e.g., number of nonreligious, nonprofit organizations), institutional health (e.g., voting rates in presidential elections and mail-back census response rates) and collective efficacy (violent crimes per 100,000 population) [33].

#### Urbanization level

The urbanization level across counties was measured using the CDC’s National Center for Health Statistics (NCHS) Rural Classification Scheme for Counties [26], a six-level urban-rural classification scheme for U.S. counties and county-equivalent entities. The six categories consist of 1) large central metropolitan, 2) large fringe metropolitan, 3) medium metropolitan, 4) small metropolitan, 5) non-metropolitan, and 6) noncore. For the purpose of data analysis in the current study, we further grouped categories 1 and 2 into “Large urban” areas, categories 3 and 4 as “Small urban” areas, and categories 5 and 6 as “Rural” areas.

### Data analysis

Given the various scales of exiting measures, we first normalized all the variables by adjusting values measured on different scales to a common scale ranged from 0 to 1 using Min-Max scaling. Bivariate analyses were performed to explore the relationship between potential predictors and their associations with vaccination rates, using Pearson correlation for continuous variables. A multivariable regression analysis was then used to examine the significance of various factors on vaccination rates. Quasi-binomial models were applied since the dependent variable is the proportion of adults fully vaccinated against COVID-19 and the proportion is conceived of as the outcome of multiple binomial trials in the quasi-binomial model. Three sequential models were employed in the regression analysis. In Model 1, only demographic characteristics were utilized. In Model 2, SDOH variables were included in addition to the demographic characteristics of Model 1. In Model 3, social capital variables (at various domains) were added to Model 2. To further explore if the impacts of social capital on vaccination rate vary by urbanization level, we stratified the final regression model by urbanization level using Model 4 (for rural areas), Model 5 (for small urban areas) and Model 6 (for large urban areas). The alpha level was set to 0.05 for regression analyses (two tailed). To assess the multicollinearity, the variance inflation factor (VIF) was calculated for Models 3 to 6. Forest plots were created based on the final regression models to visualize the associations of COVID-19 vaccination rate with all the factors including social capital variables, demographic variables, and SDOH. Data were analyzed using R (version 19.0).

## RESULTS

### Bivariate analysis

Correlations among continuous variables are shown in Table 2. Vaccination rate was associated with gender including percentage of females (r=0.05); race/ethnicity including percentage of non-Hispanic Black (r=-0.12), percentage of non-Hispanic Asian (r=0.24), percentage of non-Hispanic Native Hawaiian/ Pacific Islander (r=-0.10) and percentage of Hispanic populations (r=0.14); SDOH including percentage of living in poverty (r=-0.23), percentage of completing High school (r=0.29), median household income (r=0.38), percentage of uninsured (r=-0.33), percentage unemployed (r=0.05), and percentage of living with severe housing cost burden (r=0.15); and social capital factors including percentage of births to unmarried women (r=-0.06), number of non-religious non-profit organizations per 1000 population (r=0.12), number of religious congregation per 1000 population (r=-0.32), presidential election voting rate in 2012 & 2016 (r=0.21), and mail-back census response rate (r=0.16). In addition, vaccination rate varied by urban status (r=-0.27) with larger urban areas had a higher vaccination rate.

**Table 2.** Results of bivariate analysis.

| Potential predictors | Percentage of fully vaccination |  |
| --- | --- | --- |
|  | Pearson r | p-value |
| Percentage of female | 0.047 | <0.01 |
| Percentage of non-Hispanic Black | -0.120 | <0.01 |
| Percentage of non-Hispanic Asian | 0.244 | <0.01 |
| Percentage of non-Hispanic Native Hawaiian/Pacific Islander | -0.100 | <0.01 |
| Percentage of Hispanic | 0.138 | <0.01 |
| Percentage of living in Poverty | -0.234 | <0.01 |
| Percentage of High School Education | 0.288 | <0.01 |
| Median Household Income | 0.375 | <0.01 |
| Gini Coefficient | -0.014 | 0.42 |
| Percentage of Uninsured | -0.327 | <0.01 |
| Percentage of Unemployed | 0.049 | <0.01 |
| Percentage of Living with Severe Housing Cost Burden | 0.152 | <0.01 |
| Percentage of births to unmarried women | -0.062 | <0.01 |
| Percentage of women currently married | 0.001 | 0.96 |
| Percentage of children with single parent | -0.026 | 0.15 |
| Number of Non-religious non-profit organizations per 1000 | 0.118 | <0.01 |
| Religious congregation per 1000 | -0.321 | <0.01 |
| Presidential election voting rate, 2012 & 2016 | 0.212 | <0.01 |
| Mail-back census response rate | 0.158 | <0.01 |
| 2013_urbanization_code | -0.274 | <0.01 |
| Violent crimes per 100,000 | 0.000 | 1.0 |

### Regression analysis

Table 3 presents the hierarchical regression analysis results. Model 1 suggested that women (OR=1.41; 95% CI [1.12, 1.77]), Hispanic (OR=1.39 [1.23, 1.58]), and Non-Hispanic Asian populations (OR=22.61 [16.07, 31.83]) were associated with a higher vaccination rate. Non-Hispanic Black (OR=0.64 [0.56, 0.71]) and Non-Hispanic Native Hawaiian/ Pacific Islander (OR=0.00003 [0.000006, 0.0002]) populations were associated with a lower vaccination rate. This initial model, including only demographic factors, explained 14.5% of variance.

**Table 3.**
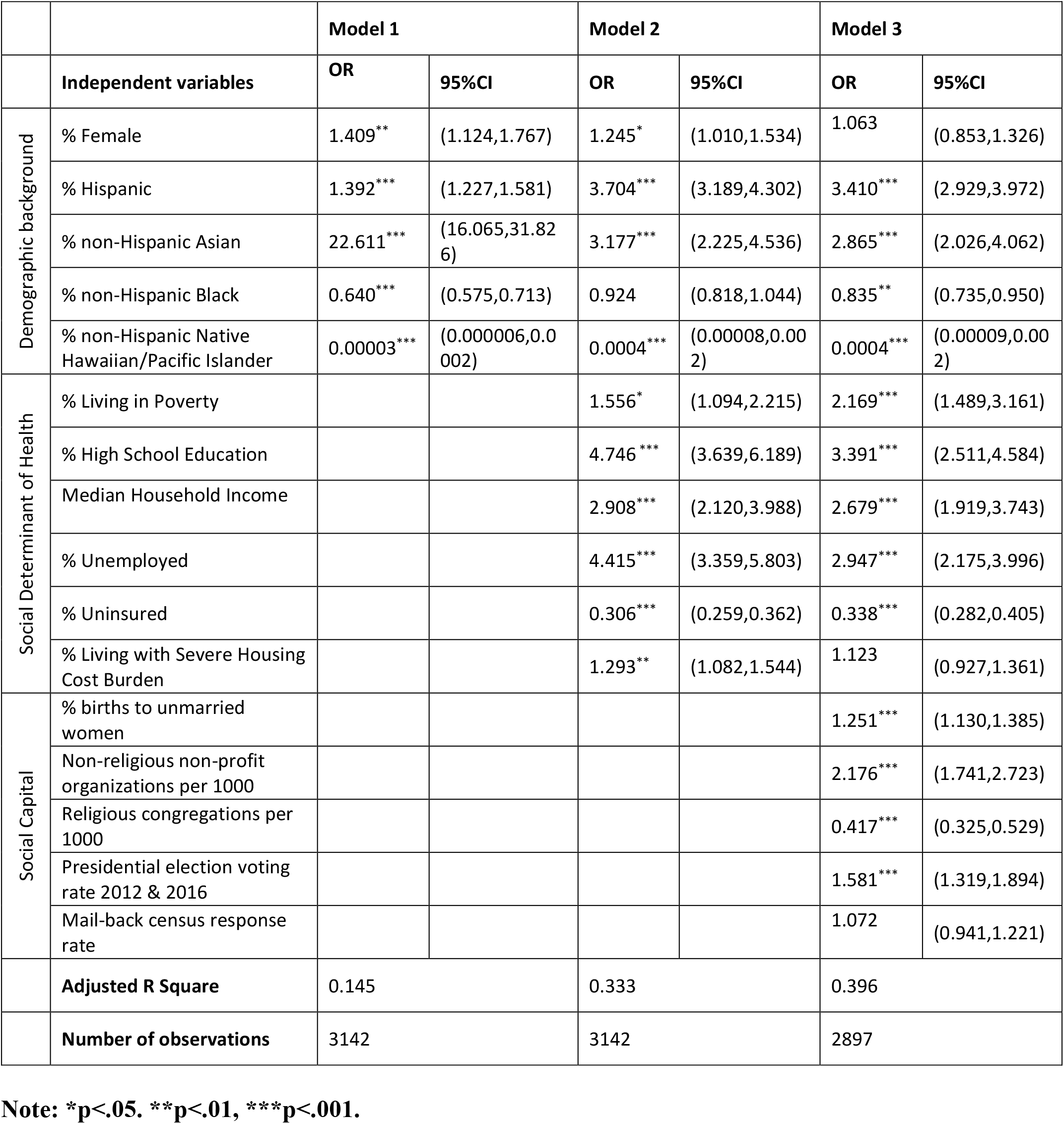
Quasi-binominal Regression results for Model 1, Model 2, and Model 3.

Model 2 showed similar results in terms of association between demographic characteristics and vaccination rate, and also suggested that SDOH that were associated with a higher vaccination rate were percentage of the population living in poverty (OR=1.56 [1.09, 2.22]), percentage completing high school (OR=4.75 [3.64, 6.19]), median household income (OR= 2.91 [2.12, 3.99]), percentage unemployed (OR=4.42 [3.36, 5.80]), and percentage living with severe housing cost burden (OR=1.29 [1.08, 1.54]). One SDOH associated with a decreased vaccination rate is the percentage of uninsured population (OR=0.31 [0.26, 0.36]). Model 2 explained 33.3% variance in vaccination rate.

The final model (i.e., Model 3) accounted for demographic factors, SDOH, and social capital factors that may influence vaccination rate. All the associations between the demographic and SDOH predictors and the vaccination rate remained significant in Model 3 except the proportion of females and the percentage living with severe housing cost burden. Among social capital factors included in the analysis, the percentage of births to unmarried women (OR=1.25 [1.13,1.39]), number of non-religious non-profit organizations per 1,000 (OR= 2.18 [1.74, 2.72]), and rates of voting in the 2012 and 2016 presidential elections (OR=1.58 [1.32, 1.89]) were associated with a higher vaccination rate, while the number of religious congregations per 1,000 (OR=0.42 [0.33, 0.53]) were associated with a lower rate. In Model 3 (i.e., the final model), variables collectively explained 39.6% variance in vaccination rate.

In summary, demographic variables explained 14.2% of the variance (Model 1) while the final model (Model 3) variables, including demographic, SDOH, and social capital variables, explained nearly 40% of the variance.

### Stratify analysis

Table 4 and Figure 1 present the results of the final regression model for rural areas (Model 4), small urban areas (Model 5), and large urban areas (Model 6). In rural areas, Hispanic populations (OR=2.63 [2.19, 3.17]) were associated with a higher vaccination rate while Non-Hispanic Native Hawaiian/ Pacific Islander populations (OR=0.002 [0.0002,0.015]) were associated with a lower rate. Model 4 suggests that the SDOH associated with higher vaccination rates included the percentage of the population living in poverty (OR=2.51 [1.55, 4.05]), percentage completing high school (OR=2.44 [1.68, 3.54]), median household income (OR=4.18 [2.37, 7.39]), and percentage unemployed (OR=4.11 [2.86, 5.91]), while the percentage uninsured (OR=0.43 [0.35, 0.54]) was associated with a lower rate. As related to social capital, the model suggests that the percentage of births to unmarried women (OR=1.23 [1.10, 1.38]), number of non-religious non-profit organizations per 1,000 (OR= 2.55 [1.91, 3.41), and rates of voting in presidential elections (OR=1.59 [1.26, 2.01]) were associated with higher vaccination rates, while the number of religious congregations per 1,000 (OR=0.40 [0.30, 0.53]) were associated with a lower rate. Model 4 has an adjusted R^2^ of 0.285.

**Table 4.**
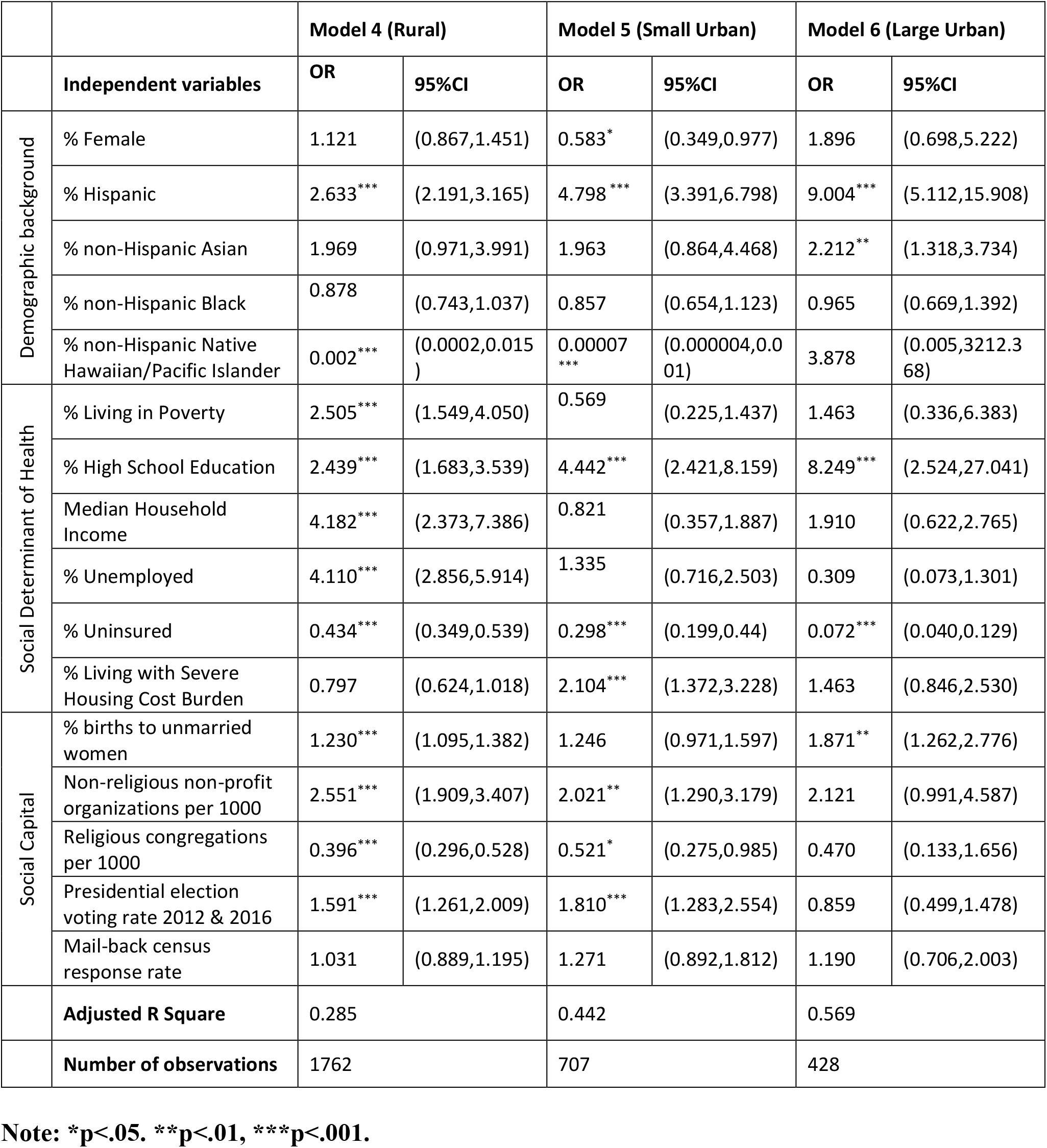
Regression results when stratified by urbanization level.

**Figure 1.**
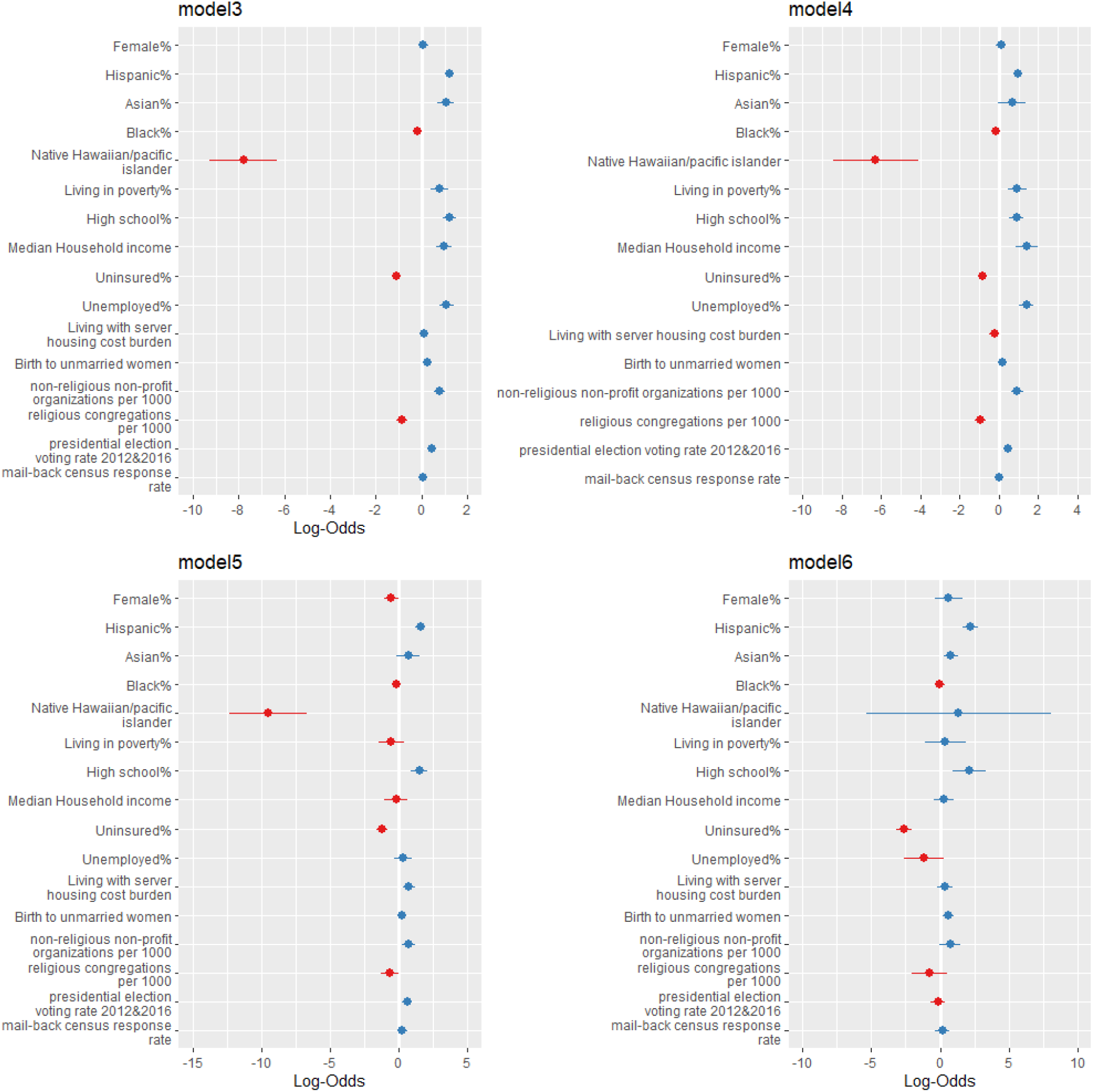
Forest plot of Model 3 to Model 6. **Notes:** Logarithm of odds ratio was used in developing the forest plots given large value of some odds ratios. We then use zero instead of one as the criteria of significance.

In small urban areas, Hispanic populations (OR=4.80 [3.39, 6.90]) were associated with higher vaccination rates while female (OR=0.58 [0.35, 0.98]) and Non-Hispanic Native Hawaiian/ Pacific Islander (OR=0.00007 [0.000004, 0.001]) populations were associated with a lower rate. As related to the SDOH, the percentage of the population completing high school (OR= 4.44 [2.42, 8.16]) and living with severe housing cost burden (OR=2.10 [1.37,3.23]) were associated with higher vaccination rates while the percentage living in poverty (OR=0.57 [0.23, 1.44]) and uninsured (OR=0.30 [0.20, 0.44]) were associated with a lower rate. Among social capital factors included in the analysis, the number of non-religious non-profit organizations per 1,000 (OR=2.02 [1.29, 3.18]) and rates of voting in presidential elections (OR=1.81 [1.28, 2.55]) were associated with higher vaccination rates, while the number of religious congregations per 1,000 (OR= 0.52 [0.28, 0.99]) were associated with a lower rate. The adjusted R^2^ of Model 5 is 0.442.

In large urban areas, Hispanic (OR=9.00 [5.11, 15.91]) and Non-Hispanic Asian (OR=2.21 [1.32, 3.73]) populations were associated with higher vaccination rates. As related to the SDOH, the percentage of the population completing high school (OR=8.25 [2.52, 27.04]) was associated with higher vaccination rates while the percentage uninsured (OR=0.07 [0.04, 0.13]) was associated with lower rates. Among the social capital factors included in the analysis, the percentage of births to unmarried women (OR=1.87 [1.26, 2.78]) was associated with higher vaccination rates. Model 6 has an adjusted R^2^ of 0.569.

The variance inflation analysis did not suggest multicollinearity (VIF values ranged from 1.03 to 7.62 across all steps).

## DISCUSSION

The current study suggests that social capital (especially its structural-level and community-level domain) contribute significantly to the disparities of vaccination uptake in the United States adjusting demographic characteristics and SDOH. The results of stratify analysis by urban-rural category show common predictors (e.g., race/ethnicity, health insurance, education attainment) of vaccine uptake but also suggest various patterns across rural, small urban and large urban areas regarding the association between COVID vaccine uptake, SDOH, and social capital factors.

The models suggest race as a strong predictor of the COVID-19 vaccination rate [34]. This is reflected as, regardless of urbanization level, higher proportions of Hispanic and Non-Hispanic Asian populations were associated with higher rates of full vaccination against COVID-19 while Non-Hispanic Native Hawaiian and Pacific Islander and Non-Hispanic Black populations were associated with lower vaccination rates. The SDOH were also found to play a significant role in likelihood of vaccination. For instance, education and median household income were found to facilitate vaccine uptake while lack of health insurance impeded uptake. As household income is related to education, level of education may influence individuals’ knowledge and perceptions regarding COVID-19 vaccines [5]. Both education level and its associated assumptions of knowledge and perceptions may contribute to vaccination hesitancy among those with limited health literacy or with mistrust in science and health agencies [35]. Additionally, a significant finding is that although the COVID-19 vaccine was administered at no cost to all people in the United States, those who are uninsured were associated with low vaccination uptake. This may be due to a lack of inclusion of uninsured individuals within existing health systems, which causes increased difficulties for their accessing health-related information and healthcare services [36].

Similarly, the relationship between social capital factors and COVID-19 vaccination rates are complex. Among factors of the community health domain, non-religious non-profit organizations, were found to be related to higher vaccination rates, likely demonstrating the presence of healthcare resources in communities and organizations that address disadvantages of the SDOH. Alternatively, religious congregations were found to be associated with a decreased vaccination rate, likely reflecting an opposition against and doubt placed on the vaccination within some religious organizations. Among factors of the institutional domain, participation in elections was associated with increased vaccination uptake. Finally, when assessing the role of family unity, percentage of births to unmarried women was associated with increased vaccination uptake. These findings, of mixed associations between social capital and vaccine uptake, reflect interesting and sophisticated decision-making dynamics demonstrating the roles of risk perceptions, social support, and community resources [11]. Generally, those with more social capital are assumed to be more likely to overcome the barriers associated with vaccination uptake through access to social networks and community support [11]. However, when individuals are living in a harsh social environment (e.g., being unemployed, being the only adult to take care of children, etc.) they may feel more vulnerable and susceptible to the impacts of the COVID-19 pandemic leading to its perception as a more severe health threat. Thus, fearing the consequences of infection and following public health guidance, individuals rightfully utilize the vaccine to protect themselves from infection.

When stratified by urbanization level, we found mixed results regarding the association between COVID-19 vaccination uptake, SDOH, and social capital factors. High school education was associated with increased vaccine uptake while lack of health insurance was found to impede vaccination uptake in all levels of urbanization considered, rural, small urban, and large urban. In rural areas, living with poverty, high median income, and unemployment status predicated a higher vaccination rate. Within small urban areas, living with severe housing cost burden was associated with increased vaccination rates while living with poverty was found to be a barrier to vaccination. In large urban areas there were no additional associations found between vaccination uptake and economic and employment status. These mixed results were unexpected and require further exploration, but these initial findings may imply that more SDOH factors affect vaccination uptake among people in rural and small urban areas compared to their counterparts in large urban areas. Further, the concrete SDOH factors may be differentiated by urban status with assumption that rural areas are more vulnerable to the disadvantages of the SDOH.

The association between vaccination uptake and social capital factors were also complicated by rural/urban status. First, more social capital variables are related to vaccine uptake rates among rural counties as compared to the small and large urban counties. This finding implies that social capital may have more of an impact on individuals’ health-related behaviors, such as vaccine uptake, in rural counties [37]. Second, among varying degrees of urbanization, vaccine uptake was associated with different social capital variables. For example, non-religious non-profit organizations and religious congregations were associated with increased and decreased vaccine uptake, respectively, in rural and small urban areas but not large urban areas. Similarly, presidential election voting rates were associated with increased vaccination uptake in both rural and small urban areas but not urban. Additionally, the percentage of births to unmarried women was associated with increased vaccination uptake in rural and large urban areas but found to have no association in small urban areas. These differentiated patterns may be explained by the varying degrees of a heterogeneous community, based on the level of urbanization, with the presence of a diversity of religions and social environments.

The current study was based on national-level dataset, which enabled us to show a comprehensive picture on disparities in vaccination rates across counties. Another strength of this work is the utilization of a hierarchical regression analysis to investigate the impact of various SDOH and social capital dimensions on vaccine uptake. Further, the stratification by urbanization level allows us to further explore how social capital affects vaccine uptake rates in different social and geospatial environments. Our study is also subject to several limitations: First, we did not include some individual-level factors such as risk perceptions and attitudes toward vaccine in data analysis, which limits our ability to explore this mechanism of social capital in affecting vaccine uptake. Second, there are missing data on some key social capital measures, which may affect the final regression results. Third, we must be cautious when interpreting the results of the stratification analysis because the sample size of model 4, 5, and 6 was quite different given the rural counties was 1589, small urban was 659, and large urban was 391. These various sample sizes result in differential statistical power.

Despite these limitations, our study demonstrates how different social capital domains may contribute to explaining the disparities of vaccine uptake based on national level data and explores how the impacts of social capital on vaccine disparities may vary by urbanization level in the United States. The study provides a new perspective to address disparities in vaccination uptake through fostering social capital within communities. Meanwhile, our findings of the stratification analysis suggest a complicated interaction between social capital and vaccine uptake across the rural-urban continuum. Further studies are needed to advance our understanding of the various associations between social capital and vaccine uptake rates by urbanization level, which have the potential to inform tailored public health intervention efforts, to enhance social capital and community resilience, and to reduce health disparities in vaccination.

## Data Availability

All the data are from openly available data sets.

https://data.cdc.gov/Vaccinations/Vaccine-Hesitancy-for-COVID-19-County-and-local-es/q9mh-h2tw

https://www.census.gov/newsroom/press-releases/2019/acs-5-year.html

https://aidsvu.org/

https://www.lee.senate.gov/scp-index

https://www.cdc.gov/nchs/data_access/urban_rural.htm

